# Physical activity traits from wrist sensors correlate with clinical status in pediatric pulmonary hypertension

**DOI:** 10.1101/2025.10.26.25338733

**Authors:** Catherine M. Avitabile, Pin-Wei Chen, Walter Faig, Babette Zemel, Jonathan A. Mitchell

## Abstract

Physical activity (PA) estimated by a wearable sensor may reflect clinical status in pediatric pulmonary hypertension (PH). Prior studies used research-grade hip-anchored sensors or commercial wrist sensors with proprietary scoring algorithms. Wrist sensors offer better acceptability in children, however, their ability to detect associations between PA and clinical characteristics is unknown. Youth 8-18 years with PH [Groups 1-4, functional class (FC) I-II] and healthy controls wore a GENEActiv accelerometer on the non-dominant wrist for 14 days. Raw acceleration data were processed using the open-source GGIR R-package. Participants completed a 6-minute walk distance (6MWD) and quality-of-life questionnaire. Muscle mass and strength were assessed by densitometry and handgrip dynamometry. Most recent cardiac testing was extracted from the medical record. Groups were compared by Fisher’s exact test, unpaired t-test, or Wilcoxon rank sum test. Multivariate regression models assessed for associations between PA and clinical metrics. Thirty PH participants (median 13.9 years, 57% female, 57% Group 1, 50% FC I) and 29 controls were included. Total PA was similar. PH participants demonstrated fewer and shorter bouts of moderate-to-vigorous PA ≥10 minutes and more time spent at lower PA intensities. In PH participants, muscle mass was positively associated with PA but 6MWD was negatively associated with PA. PA was not associated with quality-of-life. Within the PH group, worse PA traits were associated with lower FC and worse clinical testing. Wrist sensors reveal deficits in PA traits including reduced moderate-to-vigorous activity bouts and lower intensity gradients in pediatric PH.

## Introduction

Pediatric pulmonary hypertension (PH) is a rare but lethal disease with a reported prevalence of 30 per million children and 75% 5-year survival (1–5). Increased right ventricular afterload leads to right heart dysfunction, heart failure, and high risk of mortality without treatment. Only two pulmonary vasodilator medications are approved by the United States Food and Drug Administration for treatment of pediatric PH. One of the limitations to clinical trials in pediatric PH is the lack of non-invasive, easily accessible endpoints reflective of functional status in children of all ages. As dyspnea and exercise intolerance are common complaints in both children and adults with PH, the 6-minute walk distance (6MWD) is the most common functional endpoint in adult PH trials. However, the test is technically challenging or even impossible in many children due to age or developmental status (6) and typically requires travel to a dedicated pediatric PH center.

Wearable devices with an accelerometer sensor enable naturalistic estimation of physical activity (PA) in a patient’s daily life and may be more reflective of functional status than periodic, brief exercise testing. Multiple adult PH studies have demonstrated associations between lower PA measures and worse disease severity including worse quality of life, higher fatigue scores, lower 6MWD, and worse performance on cardiopulmonary exercise testing (7–13). In pediatric PH, PA measured by a hip-worn sensor correlated positively with 6MWD and negatively with functional class (reflecting worse symptoms) (14). However, children seem to be more compliant with wrist-worn sensors (15–17) which has led to interest in using wrist sensors in clinical trials with repeat activity assessments. Since commercial wrist sensors are subject to proprietary data and processing algorithms, capturing raw sensor data with a research-grade device and processing the data with open-source software is a more rigorous and reproducible approach for both observational studies and clinical trials. Standardized data processing methods are necessary to advance the use of accelerometry in PH research and clinical care. To date, no studies have estimated PA using open-source scoring software to process raw sensor data captured by a wrist-worn wearable in children with PH. Additionally, the associations between PA measured by a research-grade wrist-worn sensor and functional health status in youth with PH are unknown.

Therefore, the goals of the current study were to 1) compare PA estimates between children with PH and healthy controls and 2) to explore the associations between PA and markers of functional health status (6MWD, skeletal muscle mass and strength, quality-of-life questionnaire score, cardiac testing) in children with PH using a research-grade wrist sensor and processing the raw sensor data with open-source software to generate PA traits.

## Methods

### Study population

Ambulatory children and adolescents ages 8-18 years with World Symposium of PH Groups 1-4 and World Health Organization functional class (FC) I-II were enrolled in a cross-sectional study and compared to healthy, typically developing control participants in the same age range. For each PH participant, one control participant was enrolled matched on age in years, race, sex, and body mass index category (<5^th^ percentile=underweight, 5^th^-85^th^ percentile = normal weight, >85^th^ percentile=overweight/obese). PH participants were excluded for single ventricle physiology, moderate-severe renal or hepatic impairment, pregnancy, or significant developmental delay/inability to comply with instructions to complete the procedures. Control participants were excluded for PH, congenital or acquired heart disease, moderate-severe asthma or other pulmonary disease, musculoskeletal condition, pregnancy, moderate to severe renal or hepatic impairment, significant developmental delay/inability to comply with instructions to complete the procedures, or any chronic condition requiring routine care by a specialist. Informed consent was obtained from the parent/legal guardian of participants <18 years and of participants 18 years of age; informed assent was obtained from participants <18 years. This study was approved by the Children’s Hospital of Philadelphia Institutional Review Board (IRB #20-017827, and all data were collected between 2021 and 2024.

### Study procedures

#### PH history

PH classification, functional class, medications, brain type natriuretic peptide level, and data from most recent echocardiogram and cardiac catheterization were abstracted from the medical record. Echocardiographic data included tricuspid annular plane systolic excursion Z-score, right ventricular fractional area change, right ventricular longitudinal and free wall strain, left ventricular eccentricity index, and right ventricular/left ventricular ratio as per our comprehensive PH and right ventricular function echocardiography protocol. Right ventricular fractional area change was calculated as the end-diastolic area minus the end-systolic area divided by the end-diastolic area (18). Right ventricular longitudinal and free wall strain were measured from apical 4-chamber view using the vendor package with adjustment of the auto-tracings. By convention, strain is reported as a negative number, with greater absolute value indicating better systolic function (19). Absolute value of the strain measures was used in this study. Abnormal ventricular septal position was measured by end-systolic left ventricular eccentricity index, the ratio of the left ventricular dimension parallel to the ventricular septum to the left ventricular dimension perpendicular to the ventricular septum in parasternal short axis view at the level of the papillary muscles (18). In one report, end-systolic left ventricular eccentricity index > 1.16 best identified the presence of PH (20). Two-dimensional systolic right ventricular/left ventricular ratio was also measured in the parasternal short axis view at the level of the papillary muscles (18). Right ventricular/left ventricular ratio increases with worsening right ventricular dilation, and a value > 1 is associated with worse clinical events in PH (21).

#### Anthropometry

Weight was measured to the nearest 0.1 kg on a digital scale (Scaletronix, Skaneateles, NY). Height and sitting height were measured to the nearest 0.1 cm with a wall-mounted stadiometer (Holtain, Crymych, UK) and a sitting platform without shoes or hair adornments. Leg length was calculated as height minus sitting height. Growth variables were converted to Z-scores (standard deviation scores) as per prior studies from our team (22–26). Sex-specific Z-scores for height, weight, and body mass index relative to age were calculated using the 2000 Centers for Disease Control and Prevention growth charts (27).

#### Densitometry measures of lean mass

Body composition was measured using a Hologic Horizon fan-beam dual energy X-ray densitometer (DXA; Bedford, MA). The machine was set to array mode using Apex software. Regional and whole-body measurement of bone mineral content and density, fat mass, fat free mass, lean body mass, percent body fat, and fat distribution was obtained. Published body composition reference data were used to calculate sex- and race-specific leg lean mass Z-scores (LLMZ) relative to age (28, 29). As PH physiology is associated with impaired linear growth (30), LLMZ was further adjusted for leg length Z-score (31).

#### Upper extremity muscle strength

Bilateral forearm strength was measured with a handgrip dynamometer (Takei, Tokyo, Japan) (32). The participant stood upright with the shoulder adducted holding the dynamometer, not touching the trunk. The handle was adjusted to the hand size of the participant, and no extra body movement was allowed during testing. For each hand, 3 maximal effort trials lasting 4-5 seconds interspersed with 60-second rests were carried out. The highest value (kg) from each hand was retained for analysis. Sex-specific dominant and non-dominant handgrip Z-scores were generated using data from the 2011-2012 and 2013-2014 releases of the US National Health and Nutrition Examination Survey (NHANES) (29, 33).

#### Heath-related quality of life

Participants and parents or legal guardians completed the PedsQL (version 3.0, Cardiac Module: Pulmonary Hypertension for PH participants and standard PedsQL for control participants), a brief, practical, reliable, vaildated instrument to assess health related quality of life (34), the perception of the impact of an illness, medical therapy, or condition on a person’s ability to participate in and find satisfaction in life’s shared experiences. It assesses physical, emotional, social, and school functioning. A total summary score and physical and psychosocial health subscores are generated. Scores of 0-100 are possible with higher scores representing better quality of life.

6-minute walk distance (6MWD): Control participants performed a 6MWD at their study visit. For PH participants, the most recent standard of care 6MWD was included. As much as possible, the study visit occurred the same day as a routine clinical visit in PH participants, and the standard of care 6MWD was performed the same day as other research testing. All tests were performed according to American Thoracic Society guidelines (35). Patients walked at their own pace to cover as much distance as possible in 6 minutes along a 45-m course marked at 1-m intervals in a level hospital corridor. Heart rate and oxygen saturation were measured continuously by non-invasive pulse oximetry (Nelcor Oximax N-65, Minneapolis, MN).

#### Estimation of physical activity by wearable sensor

Participants wore a GENEActiv device on their non-dominant wrist for 14 days after the study visit. Raw sensor data were collected at a sampling rate of 30 Hz. The GGIR R-package version 3.0-0 (36) was used to process the multiple days of raw gravitational acceleration data (g/m^2^) to estimate PA parameters. First, all raw data files were auto calibrated, with the imputation processes enabled (37), and processed using the Euclidian norm minus one (ENMO) approach (38). The auto-calibration algorithm and ENMO are preprocessing steps typical in most accelerometer data. Auto-calibration estimates potential sensor calibration errors and recalibrates the activity data (37). The ENMO algorithm separates the gravitational components and the movement components, turning raw data into an easier-to-process activity intensity (38). Second, the GGIR default non-wear detection algorithm was applied (39). GGIR non-wear detection uses standard deviation and range of low acceleration values to estimate whether one is wearing the watch. Third, the Cole-Kripke sleep scoring algorithm was used with the “Heuristic algorithm looking at Distribution of Change in Z-Angle” (HDCZA) to detect sleep periods (40). HDCZA uses the estimated z-axis angle to make basic assumptions about sleep start and stop (40). This is done when there is no sleep diary to guide the sleep algorithm to accurately predict sleep onset and offset. The Cole-Kripke algorithm is one of the classic sleep algorithms to determine sleep-wake cycles. Lastly, PA levels were quantified during the remaining wake periods for each day. Since there are no PA intensity cut points validated for the PH population, we investigated three cut point settings including the most commonly used cut point in GGIR (GGIR default) and two that partially matched with the study age group and device/wear location: Schaefer 2014 (children ages 6-11 years, GENEActiv wrist) (41) and Hildebrand 2014 (adults and children ages 7-11 years, ActiGraph hip/GENEActiv wrist) (42). When examining the face validity of the moderate-to-vigorous PA (MVPA) outputs, daily estimates were either much too low (median 0 minutes with no range with Hildebrand) or too high (median 25-30 minutes with Schaefer) based on prior work in the PH population. Therefore, even if these settings matched with the specific age range/device, the face validity did not support using them when studying pediatric PH participants and comparing them to a group of relatively healthy individuals of the same age range. Based on this, we used the most common GGIR default cut points which demonstrated better face validity. Three general filters were used to screen out days with extreme values: 1) daytime data ≤ 8 hours; 2) daytime non-wear percentage ≥25%; and 3) daytime accumulation >24 hours. These filters serve to reduce the number of errors from GGIR estimations.

The default GGIR cut points were used to estimate time spent in sedentary behavior and PA intensities (minutes per day): sedentary, <30 gravitational units; light physical activity, 30-100 gravitational units; moderate physical activity, 101-400 gravitational units; and vigorous physical activity, ≥400 gravitational units. Since typical arm movements may inflate PA estimates, bouts of physical activity were also estimated. For MVPA, bouts were reported at 1-, 5-, and 10-minute intervals; for light PA and sedentary behavior, bouts were calculated at 1-, 10-, 30-minute intervals. Note, a bout was defined as a timed interval of continuous movement, of which ≥80% of the time interval is spent in sedentary behavior or at a specific PA intensity.

GGIR was also used to generate the PA intensity gradient (43). The intensity gradient sorts the participant’s full spectrum of PA data into intensity-based bins and uses the log–log slope of the intensity and time accumulated at each intensity to quantify the distribution of the individual’s PA intensity. The intensity gradient is a cut point-independent metric. The slope of the intensity gradient is negative to reflect less time spent at higher intensities. A steeper, more negative, slope with a higher constant (y-intercept) indicates a steep drop in time accumulated with increasing intensity and an uneven distribution of time spent across all intensities (e.g., more time at lower intensities); a shallower slope indicates a more even distribution of time spent at all PA intensities (e.g., more time at higher intensities) (43).

### Statistical analyses

Standard descriptive statistics [mean ± standard deviation or median (interquartile range)] were used to summarize demographics, clinical characteristics, LLMZ, handgrip Z-scores, quality of life scores, and PA. All days of PA data were analyzed and daily averages over the observation period are presented. Comparisons between PH and control participants were performed by Fisher’s exact test for categorical variables and unpaired t-test or Wilcoxon rank sum test (if not normally distributed) for continuous variables. Multivariate regression models were created to assess for associations between PA and demographic characteristics (including age, sex, and height Z-score), LLMZ, handgrip Z-score, quality of life scores in PH and control participants. Univariate regression was used to assess for associations between PA and PH disease-specific characteristics in PH participants. 6MWD was scaled by 100-meter change. Variable-by-PH status interaction terms were included to consider differential associations by PH status. Analyses were conducted in SAS with two-sided tests of hypotheses and a p*-*value <0.05 as the criterion for clinical significance.

## Results

Thirty PH participants and 30 healthy control participants were enrolled. One healthy control participant was excluded from analysis due to ≤ 8 hours of daytime data on all observation days. Age, sex, race, and ethnicity compositions were similar between the PH and control participants (Table 1). Height and weight Z-scores, LLMZ, handgrip Z-scores, and 6MWD were lower in PH participants compared to control participants (Table 1). PedsQL total scores were comparable between PH and control participants (Table 1).

**Table 1.** Characteristics of PH and healthy control participants. ^1^In PH participants, the median (interquartile range) time interval between 6MWD and start of PA assessment was 0 (0,0) days. Data expressed as count (%) or median (interquartile range). PH, pulmonary hypertension; BMI, body mass index; 6MWD, 6-minute walk distance

|  | **PH (n=30)** | **Controls (n=29)** | **p value** |
| --- | --- | --- | --- |
| Age, years | 13.9 (9.6, 15.8) | 14.2 (10.8, 15.1) | 0.89 |
| Female, % | 17 (57) | 17 (59) | 0.88 |
| Black race, % | 8 (27) | 6 (21) | 0.59 |
| Hispanic or Latino ethnicity, % | 2 (7) | 3 (10) | 0.61 |
| Height Z-score | -0.68 (-1.18, 0.00) | -0.15 (-0.63, 0.56) | 0.02 |
| Weight Z-score | -0.57 (-1.21, 0.25) | 0.10 (-0.50, 0.52) | 0.04 |
| BMI Z-score | -0.33 (-1.11, 0.66) | 0.32 (-0.62, 0.69) | 0.19 |
| Leg lean mass Z-score | -1.65 (-2.19, -0.87) | -0.87 (-1.13, -0.39) | <0.01 |
| Dominant handgrip Z-score | -1.50 (-2.23, -0.59) | -0.55 (-0.98, 0.13) | <0.01 |
| Non-dominant handgrip Z-score | -2.09 (-2.69, -0.77) | -0.75 (-1.27, 0.02) | <0.01 |
| 6MWD<sup>1</sup>, m | 594 (500, 634) | 694 (633, 731) | <0.01 |
| PedsQL total score | 74.2 (67.6, 82.4) | 82.6 (70.9, 90.2) | 0.08 |

Clinical characteristics of the PH participants are detailed in Table 2. Most participants were World Symposium of PH Group 1 or 3. Participants were evenly split between FC I and II. Most were treated with dual enteral vasodilator therapy at the time of the study, however 23% were on triple therapy. Median BNP was low. Quantitative echocardiographic assessment reflected normal to mildly diminished right ventricular systolic function without significant right ventricular dilation on average. Summary cardiac catheterization data for the cohort were consistent with mild elevation in pulmonary artery pressure and pulmonary vascular resistance and normal cardiac index.

**Table 2.** PH-specific clinical characteristics.

|  | PH (N=30) |
| --- | --- |
| World Symposium of PH classification |  |
| 1 | 17 (57) |
| 2 | 2 (7) |
| 3 | 10 (33) |
| 4 | 1 (3) |
| World Health Organization functional class |  |
| I | 15 (50) |
| II | 15 (50) |
| Number of PH medications |  |
| ≤ 2 | 23 (77) |
| 3 | 7 (23) |
| Brain type natriuretic peptide level, *pg/mL* | 32.2 (11.9, 55.3) |
| Echocardiographic data |  |
| Interval between echocardiographic data and start of PA assessment, *d* | 0 (0, 6) |
| TAPSE Z-score | -2.3 (-5.1, 0.2) |
| RV fractional area change, % | 37.3 (30.1, 41.2) |
| RV longitudinal strain, % | 21.6 (14.6, 24.8) |
| RV free wall strain, % | 24.0 (17.1, 28.8) |
| RV/LV ratio | 0.8 (0.7, 1.0) |
| LV eccentricity index | 1.1 (1.0, 1.3) |
| Cardiac catheterization data |  |
| Interval between cardiac catheterization data and start of PA assessment, *m* | 21 (8, 38) |
| Mean pulmonary artery pressure, *mm Hg* | 27 (20, 35) |
| Cardiac index, *L/min/m<sup>2</sup>* | 3.9 (3.5, 4.0) |
| PVRi, *wood unit\*m<sup>2</sup>* | 4.5 (2.9, 6.1) |

Data expressed as count (%) or median (interquartile range).
PH, pulmonary hypertension; TAPSE, tricuspid annular plane systolic excursion; RV, right ventricular; LV, left ventricular; PVRi, indexed pulmonary vascular resistance

PA characteristics are displayed in Table 3. The number of valid wear days was similar between the groups. Total PA time and time by intensity level (sedentary, light PA, and MVPA) did not differ between the groups. However, there were differences in bouts of MVPA. PH participants demonstrated fewer [median 0.5 (IQR 0.13, 1.5) in PH vs. 1.6 (0.9, 3.8) in controls] and shorter [7.7 (1.3, 16.6) in PH vs. 24.8 (11.0, 44.5) in controls] bouts of MVPA ≥10 minutes. PH participants, on average, engaged in 15 minutes less MVPA when defined in bouts ≥10 minutes. PH participants also demonstrated greater number [7 (4, 9) in PH vs. 4 (2, 7) in controls] and higher percentage [69 (38, 88) in PH vs. 33 (15, 62) in controls] of days with zero bouts of MVPA ≥10 minutes achieved.

**Table 3.** Physical activity characteristics summarized over observation period.

|  | **PH (n=30)** | **Controls (n=29)** | **p value** |
| --- | --- | --- | --- |
| Number of valid wear days | 11 (8, 13) | 13 (11, 13) | 0.20 |
| Total activity time, *min/day* | 980 (911, 1037) | 964 (936, 1005) | 0.83 |
| Total sedentary time, *min/day* | 615 (537, 699) | 561 (510, 707) | 0.47 |
| Total light activity time, *min/day* | 225 (204, 275) | 234 (194, 272) | 0.73 |
| Total MVPA time, *min/day* | 115 (67, 143) | 127 (99, 180) | 0.14 |
| MVPA bouts 5-10 minutes |  |  |  |
| Number per day | 1.2 (0.7, 3.7) | 2 (1.2, 3.4) | 0.14 |
| Duration, *min* | 6.3 (2.6, 15.6) | 10.6 (5.8, 18.2) | 0.16 |
| Number of days with zero bouts | 4 (1, 6) | 3 (1, 5) | 0.38 |
| Percent days with zero bouts | 36 (9, 69) | 23 (8, 50) | 0.40 |
| MVPA bouts ≥10 min |  |  |  |
| Number per day | 0.5 (0.13, 1.5) | 1.6 (0.9, 3.8) | <0.01 |
| Duration, *min* | 7.7 (1.3, 16.6) | 24.8 (11.0, 44.5) | <0.01 |
| Number of days with zero bouts | 7 (4, 9) | 4 (2, 7) | 0.02 |
| Percent days with zero bouts | 69 (38, 88) | 33 (15, 62) | <0.01 |
| Intensity gradient |  |  |  |
| Slope | -2.25 (-2.42, -2.09) | -2.11 (-2.32, -1.93) | 0.05 |
| Intercept | 13.3 (12.7, 14.0) | 12.7 (12.0, 13.7) | 0.08 |

Data expressed as count (%) or median (interquartile range).
PH, pulmonary hypertension; MVPA moderate to vigorous physical activity

The slope of the intensity gradient was more negative in PH participants compared to control participants (Table 3). This finding reflects a steeper slope of the intensity gradient and more time spent at lower intensities. The intercept of the intensity gradient was comparable between PH and control participants.

When adjusted for age, sex, and height Z-score, PH status remained associated with shorter MVPA bouts ≥10 minutes (p<0.01) and steeper slope of the intensity gradient (p<0.01) compared to control participants. PH status nearly reached statistical significance for number of MVPA bouts ≥10 minutes (p=0.05). PH status was not significant for other PA traits when adjusted for age, sex, and height Z-score.

Multivariate regression models for associations between PA and participant characteristics in PH and healthy controls are displayed in Table 4. Each model included a single PA trait. In comparing PH participants to healthy controls, the interaction analyses revealed several instances where PH status changed the impact of an exposure on the PA traits including unbouted MVPA, MVPA bouts ≥10 minutes, and slope of the intensity gradient. On average, unbouted MVPA was lower in female participants compared to male participants. However, PH status attenuated the lower MVPA in females compared to males (Table 4A, model #1). For example, compared to control males, average daily unbouted MVPA was 12.5 minutes less in control females, 14.5 minutes less in PH males, and 9.1 minutes less in PH females. Sex differences in MVPA were less pronounced in PH participants compared to control participants. Similarly, on average, unbouted MVPA was lower in Black participants compared to non-Black participants. Again, PH status attenuated the lower MVPA in Black participants compared to non-Black participants (Table 4A, model #2). Compared to non-Black control participants, average daily unbouted MVPA was 16 minutes less in Black controls, 6.9 minutes less in non-Black PH participants, and 7.1 minutes less in Black PH participants. Racial differences in MVPA were less pronounced in PH participants compared to control participants.

**Table 4.** General linear regression models for the associations between PA and participant characteristics in PH and healthy controls. Multivariate models with A) unbouted MVPA as dependent variable, B) slope of the intensity gradient as dependent variable, C) number of MVPA bouts ≥10 minutes as dependent variable, and D) duration of MVPA bouts ≥10 minutes as dependent variable. Each model includes a single PA trait.

| Model # | Dependent variable | Independent variables | Estimate | SE | P value |
| --- | --- | --- | --- | --- | --- |
| 1 | Unbouted MVPA | Female | -12.52 | 3.29 | <0.01 |
|  |  | PH | -14.51 | 3.65 | <0.01 |
|  |  | Female*PH | 17.89 | 4.73 | <0.01 |
| 2 | Unbouted MVPA | Black race | -15.95 | 4.17 | <0.01 |
|  |  | PH | -6.90 | 2.62 | 0.01 |
|  |  | Black *PH | 15.75 | 5.74 | <0.01 |

B)
| Model # | Dependent variable | Independent variables | Estimate | SE | P value |
| --- | --- | --- | --- | --- | --- |
| 1 | Slope of the intensity gradient | Female | -0.16 | 0.03 | <0.01 |
|  |  | PH | -0.23 | 0.03 | <0.01 |
|  |  | Female*PH | 0.11 | 0.05 | 0.02 |
| 2 | Slope of the intensity gradient | Black race | -0.24 | 0.04 | <0.01 |
|  |  | PH | -0.19 | 0.02 | <0.01 |
|  |  | Black*PH | 0.15 | 0.05 | <0.01 |
| 3 | Slope of the intensity gradient | LLMZ | -0.08 | 0.03 | <0.01 |
|  |  | PH | -0.03 | 0.04 | 0.45 |
|  |  | LLMZ*PH | 0.12 | 0.03 | <0.01 |

C)
| Model # | Dependent variable | Independent variables | Estimate | SE | P value |
| --- | --- | --- | --- | --- | --- |
| 1 | Number of MVPA bouts $\geq 10$ minutes | 6MWD<sup>1</sup> | 0.34 | 0.10 | <0.01 |
|  |  | PH | 2.43 | 0.83 | <0.01 |
|  |  | 6MWD*PH | -0.40 | 0.13 | <0.01 |

<sup>1</sup>6MWD scaled by 100 m
SE, standard error; MVPA, moderate to vigorous physical activity; PH, pulmonary hypertension; LLMZ, leg lean mass Z-score; 6MWD, 6-minute walk distance

Similar findings were seen with the slope of the intensity gradient (Table 4B). On average, the slope of the intensity gradient was more negative (steeper = more time spent at lower intensities) in female participants compared to male participants and Black participants compared to non-Black participants. However, PH status attenuated these sex and race group differences and resulted in a shallower slope of the intensity gradient (Table 4B, model #1 and #2).

In PH participants, LLMZ attenuated the negative slope of the intensity gradient (Table 4B, model #3) and was positively associated with longer MVPA bouts ≥ 10 minutes (Table 4D, model #1). However, while 6MWD was positively associated with PA overall, the association was negative in PH participants. In PH participants, 6MWD was negatively associated with the number (Table 4C, model #1) and duration (Table 4D, model #2) of MVPA bouts ≥ 10 minutes. For each 100-meter increase in 6MWD, the number of daily MVPA bouts ≥ 10 minutes decreased by 0.4 and the duration of MVPA bouts ≥ 10 minutes decreased by 0.37 minute. This negative association persisted after controlling for age, gender, and height Z-score.

Univariate regression models within the PH group are displayed in Table 5. Each model included one PA trait and one PH characteristic. Associations were identified between individual PA traits (unbouted MVPA, duration of MVPA bouts ≥10 min, and slope of the intensity gradient) and indicators of PH severity [right heart function measured by right ventricular fractional area change on echocardiogram, cardiac catheterization hemodynamics (mean pulmonary artery pressure, cardiac index, indexed pulmonary vascular resistance), and functional class (symptom severity)]. Associations were consistent with clinical severity: higher right ventricular fractional area change (better heart function) was associated with better PA traits while higher mean pulmonary artery pressure and indexed pulmonary vascular resistance (worse hemodynamics) and higher functional class (worse symptomatology) were associated with worse PA traits. PA traits discriminated by functional class (Figure). There were no associations between PA and brain type natriuretic peptide level or other echocardiographic measures right heart function within the PH group.

**Table 5.** General linear regression models for the associations between PA and PH characteristics. Univariable models with A) unbouted MVPA as dependent variable, B) duration of MVPA bouts ≥10 minutes and C) slope of the intensity gradient as dependent variable. Each model includes one PA trait and one PH characteristic.

| Dependent variable | Model # | Independent variable | Estimate | SE | p value |
| --- | --- | --- | --- | --- | --- |
| Unbouted MVPA | 1 | RV fractional area change | 1.1 | 0.24 | <0.01 |
|  | 2 | Mean pulmonary artery pressure | -0.29 | 0.14 | 0.05 |
|  | 3 | Cardiac index | 5.52 | 1.46 | <0.01 |
|  | 4 | PVRi | -1.37 | 0.58 | 0.03 |
|  | 5 | Functional class (II vs. I) | -14.9 | 3.5 | <0.01 |

B)
| Dependent variable | Model # | Independent variable | Estimate | SE | p value |
| --- | --- | --- | --- | --- | --- |
| Duration of MVPA bouts $\geq 10$ min | 1 | RV fractional area change | 0.01 | 0.01 | 0.24 |
|  | 2 | Mean pulmonary artery pressure | 0.00 | 0.01 | 0.99 |
|  | 3 | Cardiac index | 0.04 | 0.05 | 0.44 |
|  | 4 | PVRi | 0.00 | 0.03 | 0.99 |
|  | 5 | Functional class (II vs. I) | -0.44 | 0.16 | <0.01 |

## Discussion

This is the first study to process raw acceleration data from a research-grade wrist sensor to derive estimates of PA in pediatric PH patients and demonstrate deficits in PA levels compared to healthy peers and in relation to PH disease severity. We observed that PH participants demonstrated fewer and shorter daily MVPA bouts greater than 10 minutes and steeper decline in the intensity gradient indicating an uneven distribution of time spent across all PA intensities and more time spent at lower intensities. In PH participants, LLMZ attenuated the negative slope of the intensity gradient and was positively associated with PA bouts suggesting an important role for skeletal muscle in these patients. Within the PH group, less time spent in MVPA (overall and in bouts) and steeper decline in the intensity gradient were associated with higher PH severity. These findings indicate that bouted MVPA estimates and intensity gradient estimates may have utility in pediatric PH. Unbouted MVPA estimates from wrist-worn devices are likely less reliable due to brief motion artifacts such as short arm movements which are not reflective of physical exertion. Uncovering differences in bouted PA may reflect a fundamental lack of endurance in PH and lead to new clinical trial outcomes.

Despite many medication trials in adult PH, there have been very few in children (44–46). One major limitation to clinical trials is the lack of relevant trial endpoints reflective of hemodynamics or functional status in children in whom standard exercise testing is impractical, unreliable, or not reproducible (6). Wearable sensors facilitate observation of patients at home without travel to a sophisticated testing center. MVPA estimated by a research-grade hip-worn sensor and analyzed in the vendor’s software platform was associated with longer 6MWD and lower functional class (14). However, greater acceptability of and compliance with wrist worn devices in children (16) has generated enthusiasm for using these sensors in clinical care and clinical trials (47). In a multi-center observational study performed within the North American Pediatric Pulmonary Hypertension Network (PPHNet), it was feasible to measure PA in even the youngest pediatric PH patients using both a research-grade hip sensor and a commercial wrist-worn sensor (48).

In contrast to the raw acceleration data obtained from research-grade sensors (49, 50), commercial wrist sensors use proprietary software and non-transparent data processing algorithms to convert raw acceleration data to activity volume and intensity. Raw data are not available to the investigator, limiting data analyses. Algorithms can change or devices can be removed from the market without notice, disrupting research and clinical care. Additionally, U.S. federal legislation – the Children’s Online Privacy Protection Act – limits the upload of activity data to commercial websites or online services, such as commercial activity tracker platforms, in kids younger than 13 years. Even with parent accounts, computing expertise is needed to draw data from commercial cloud storage into a research database. Ours is the first pediatric PH study to measure PA with the research-grade GENEActiv wrist sensor, analyze the raw PA data, and demonstrate associations between wrist-derived PA and clinical parameters. The raw data can be re-analyzed as new open-source methodologies become available, increasing rigor and reproducibility.

When using research-grade devices, investigators apply published “cut points” to translate activity counts to PA intensity levels [e.g. sedentary, light, MVPA] (51, 52). In other pediatric PH studies, this has been done within the ActiGraph vendor’s software (14, 48). Various cut points are published from distinct, small metabolic studies of healthy people within limited age ranges (53–55). This limits comparisons among studies with different conditions, ages, devices, or wear locations. PA data from prior studies in pediatric PH have been analyzed using hip cut points (14, 56). Thresholds of activity counts and MVPA that correspond to changes in clinical parameters in children with PH have been published for hip worn ActiGraph sensors using hip-derived cut points (57), however, it is critical to note that the best cut-point for estimation of PA intensity from wrist-worn devices in children is unknown. It is important that hip thresholds are not applied to wrist acceleration data since wrist acceleration is greater than hip acceleration with certain activities (16, 49).

Cut points to estimate time spent in PA intensities, specific to wrist acceleration data, are available in GGIR. In our study, there were no differences in total PA, PA of various intensity levels (sedentary, light, MVPA), or unbouted PA in PH participants compared to healthy control participants. However, when bouts of MVPA greater than 10 minutes were examined, PH participants demonstrated fewer and shorter bouts. They had more days without any 10-minute bouted activity compared to healthy control children. These findings suggest that total PA intensity and volume measured by a wrist sensor may be contaminated by brief arm movements not reflective of full body PA. Measuring MVPA bouts may be more reflective of sustained exertion. Our study also highlights the potential importance of the intensity gradient, a cut point-independent metric in pediatric PH. PH participants demonstrated steeper (more negative) slope of the intensity gradient reflecting an uneven distribution of time spent across all intensities with more time at lower intensities and less time at higher intensities. The impact of PH status on duration of MVPA bouts ≥10 min and slope of the intensity gradient persisted after controlling for age, sex, and height Z-score. These findings may reveal unique limitations in endurance in pediatric PH and may be new targets for clinical trials. Both known (MVPA) and new (MVPA bouts and intensity gradient) traits associated with PH severity by echocardiogram, cardiac catheterization, and functional classification in the expected direction. Even in these “stable,” prevalent PH patients with relatively “reassuring” echocardiographic and hemodynamic assessment, PA was different than control patients and correlated with clinical severity. The findings suggest that PA estimated by a research-grade wrist sensor may have potential as a functional endpoint in pediatric PH. It is key to study these individual wrist-derived PA traits in larger studies to evaluate their suitability as outcomes in clinical trials.

PH status attenuated sex- and race-group differences in wrist-derived PA metrics, indicating that PH physiology has a greater effect on PA than other demographic characteristics. However, the effect of LLMZ on some PA traits was greater in PH participants compared to healthy control participants. This finding is consistent with our prior data demonstrating LLMZ deficits (skeletal muscle deficits) in association with poor exercise performance in youth with PH (24, 25). Interventions to augment skeletal muscle deficits may improve PA and exercise performance in children with PH, as has been demonstrated in several adult PH exercise trials (58–60). Interestingly, the effect of 6MWD on some PA traits was less in PH participants compared to healthy control participants. These findings should be explored in larger studies with longitudinal data to determine if there is an association between wrist-measured PA and 6MWD in children with PH. PA estimation by a wearable sensor may augment home monitoring rather than one-time evaluation in a testing environment, especially in individuals who have difficulty performing standard exercise testing.

Our study has limitations. First, the sample is relatively small and originates from a single center PH program. The study included patients from World Symposium of PH Groups 1-4 which may contribute to heterogeneity of results. A larger, multi-center study may identify activity profiles specific to PH Groups (i.e. Group 1 vs. Group 3). Participants were limited to those in functional classes I and II. More profound differences in PA traits and stronger associations with clinical characteristics may be evident with a broader sample including PH patients with more advanced disease. Secondly, the study included patients 8-18 years of age. Future studies will apply these methods to wrist sensors in younger children (≤8 years) who universally cannot perform the 6MWD and other exercise testing and have the most urgent need for age-appropriate endpoints. Third, this study included prevalent PH patients, often several years from presentation and diagnosis, examined in a cross-sectional design. This study design does not allow us to predict clinical status from PA traits. However, it is notable that despite our aggressive treatment, well-managed disease, and relatively “reassuring” cardiac testing, these patients have reduced PA compared to controls and their PA parameters correlate with clinical severity. These findings should be testing in longitudinal studies including incident patients. Finally, the PA metrics in this study (other than intensity gradient) are cut point dependent. Future studies will explore device- and cut point-independent metrics including monitor independent movement summary (MIMS) units and PA percentiles derived from national representative PA data (61).

In conclusion, PH participants engaged in less PA compared to healthy controls as indicated by fewer and shorter MVPA bouts ≥10 minutes and steeper slope of the intensity gradient estimated by a wrist-worn accelerometer and an open-source scoring algorithm. PA traits also associated with disease severity in PH participants. With further research, these traits may reveal important endurance deficits in this population and could be potential targets for clinical trials.

## Data Availability

All data produced in the present study are available upon reasonable request to the authors.

## Acknowledgments

We express our appreciation to all the participants in the study as well as the various study coordinators who supported the study.

## Figure legend

PA traits including unbouted MVPA, number and duration of MVPA bouts, and intensity gradient discriminate by functional class (Blue=Functional Class I, Red=Functional Class II).

## Ethical approval

The study was approved by Children’s Hospital Institutional Review Board, protocol #20-017827.

## Guarantor

Catherine M. Avitabile MD

## Contributorship

CMA designed and executed the study and wrote the manuscript.

P-WC performed analyses of sensor data, contributed to data interpretation, and edited the manuscript.

WF performed statistical analyses and edited the manuscript.

BSZ and JAM provided senior mentorship in study design and execution, contributed to data interpretation, and edited the manuscript.

